# Prevalence, associated factors, and pharmacotherapy of hypertensive disorders among parturients in Bono Region of Ghana: An analytical cross-sectional study

**DOI:** 10.1101/2025.05.18.25327871

**Authors:** Francis Fordjour, Edward Tieru Dassah, Jonathan Boakye-Yiadom, Kwadwo Addai-Darko, Bernard Okyere, Kwame Ohene Buabeng

## Abstract

**Background:** The burden of hypertensive disorders among parturients in remote locations of Ghana remains understudied, considering existing data. We investigated the prevalence, associated factors, and treatment guideline’s adherence for maternal hypertensive disorders among parturients in a peripheral region.

**Method:** In an analytical cross-sectional study, data of all parturients with hypertensive disorders were extracted from labor registers of nine public hospitals in the Bono Region of Ghana from January to December 2021. Additionally, guidelines for administering magnesium sulfate and antihypertensives to these patients were examined. Chi-square and multivariable binomial regression analyses were used to explore associations between independent and dependent variables. P ≤ 0.05 was considered statistically significant.

**Results:** Hypertensive pregnancies were 711 out of 16,206 deliveries, with a prevalence of 4.4%. Non-severe pre-eclampsia (30.5%) and gestational hypertension (28.0%) were the most frequent disorders. Eclampsia (6.2%) and superimposed pre-eclampsia (1.7%) were less frequent. Maternal age, 15-25 years (cOR = 2.43; 95% CI = 1.57-3.75; p < 0.001), unemployment (cOR = 2.14; 95% CI = 1.29-3.53; p = 0.001), primigravida (cOR = 2.88; 95% CI = 1.80-4.62; p < 0.001), and primiparity (cOR = 2.39; 95% CI = 1.44-3.96; p < 0.001) were significantly associated with pre- eclampsia/eclampsia. After adjustment for confounders, primiparity remained a borderline significant predictor (aOR = 1.83; 95% CI= 0.97-3.46; p = 0.05). Oral nifedipine (30mg) and intravenous hydralazine were the primary medications for pregnant hypertensives. Magnesium sulfate was universally administered by the Pritchard procedure, but product concentrations supplied by pharmacists for intramuscular application varied minimally.

**Conclusion:** The prevalence of hypertensive disorders among parturients was 4.4%. Young parturient age, unemployment, primigravida, and primiparity were the predictors. Magnesium sulfate protocols for managing pre-eclampsia/eclampsia cases followed a standard regimen. Standardizing the concentration of magnesium sulfate solutions for pre-eclampsia/eclampsia could optimize intramuscular dosing under the Pritchard regimen, improving treatment consistency.

## Introduction

Hypertensive disorders in pregnancy (HDPs) continue to hinder the course of many pregnancies resulting in abrupt interventions, complications and mortalities with a potential of undermining target 3.1 of the Sustainable Development Goals (SDG) (1). The spectrum of HDPs, which varies from chronic, gestational, pre-eclampsia/eclampsia and superimposed pre-eclampsia, impact women health worldwide and have had significant repercussions for low-income nations (2,3) These disorders have become topical recently due to the rise in cases that have been documented and the unfavorable consequences they have in certain nations (4,5). The incidence of HDPs has increased by 10.92% globally over the last three decades, having risen from 16.30 million in 1990 to 18.08 million in 2019 (4).

Given that almost a third of affected women nearly die, particularly in nations with limited resources, this rising trend is extremely concerning and unnecessarily causes panic among parturients and medical personnel (6). Hypertensive disorders of pregnancy together with hemorrhage, sepsis, and unsafe abortions are major contributors to the maternal death ratio in low-income countries (7,8), surpassing the SDGs’ estimate by more than six-fold (1).

According to World Health Organization (WHO) data (9), Ghana still has a high maternal mortality ratio (263/100,000), and the country’s rising HDP burden adds to these deaths (10). Studies available on the subject almost a decade ago suggest that the incidence of HDPs in Ghana’s Upper West and Greater Accra regions is less than 10% (11). However, the prevalence was high when explored in 2019 and 2017 at the Komfo Anokye Teaching Hospital (KATH) in Kumasi, Ghana, and the Korle-Bu Teaching Hospital (KBTH) in Accra respectively (12,13). In Ghana, it is reported that over a third of women with HDPs experience ‘near-death’ episodes; the ratio of these incidents to death is approximately 12:1 (14). Despite the negative impact, it appears that the majority of research looking for HDPs in Ghana concentrate on tertiary hospitals, with little information published from lower-tier facilities, particularly those located in the rural and smaller administrative regions. There is a need to close this information gap because it is argued that the unavailability of published statistics on HDPs in smaller regions significantly affects regional and global estimations which may mislead policies (15).

Many maternal deaths, including those caused by HDPs in sub-Saharan Africa, are listed by the WHO as avoidable, implicating care plans offered to maternal hypertensives in the sub-region (8). The essential ideas that underpin the management of HDPs worldwide include early antenatal screening for case identification, antihypertensive therapy, the administration of anticonvulsants, and timely baby delivery. However, the way these concepts are applied may vary based on the resources, patient characteristics, and skill set that are available in a given geographical area. Inconsistencies were found in post-implementation studies involving the administration of magnesium sulfate (MgSO_4_) to women who were eclamptic. Some nations lacked the necessary skills to safely administer the product, while others had difficulties with product accessibility. Furthermore, some institutions lacked treatment-guided protocols for MgSO_4_ application, which hindered the safe use of the anticonvulsant in nations that have high rates of HDPs (16,17). Ghana’s standard treatment guideline (STG) incorporates all of the fundamental HDP management principles; nevertheless, there aren’t many post-implementation studies assessing these in healthcare institutions. In this study, we reviewed practice guidelines on the use of MgSO_4_ and antihypertensives for HDPs as well as the prevalence and factors associated with these disorders among parturients in the Bono Region.

## Materials and method

### Study design and setting

An analytical cross-sectional study design was employed in which obstetric records, particularly labor registers, and institutional treatment protocols for HDPs were monitored retrospectively for data in nine public hospitals of the region. The Sunyani Municipal, Sunyani SDA, Bono Regional, Berekum Holy Family, Dormaa Presbyterian, Drobo St. Mary’s Catholic, Sampa Government, Tain District, and the Wenchi Methodist Hospitals were the purposefully chosen research facilities. These hospitals were selected because they provided important obstetric services in eight of the region’s twelve administrative districts and were the most established and resourced facilities. Health facilities in four districts were at the time of data collection not included due to their status as health centers which could not fully manage HDPs. The nine hospitals had a maternal bed capacity of 384 and conducted about 1800 deliveries annually.

The Bono Region is located on Ghana’s middle belt zone, sharing an international boundary to the west with the Republic of Cote d’Ivoire and to the north and south with Ghana’s Savannah and Ashanti regions, respectively. With a population of roughly 1,208,649, the region occupies an area of 11,113 km^2^ (18). Its residents are primarily famers and traders.

### Study population and Sample

All pregnant women who were registered and delivered at the public hospitals in Bono Region of Ghana between 3^rd^ January 2021, and 31^st^ December 2021, were considered. Overall, 16,206 deliveries were conducted within the period of which 711 women presented with various HDPs. All 711 parturients diagnosed with HDPs during the study period were included in the study. Women with incomplete records during the review were excluded.

### Data collection

Data of parturients was extracted from the labor registers of the hospitals using a designed template. Women’s age, level of education, occupation, parity, gestational ages at labor, and the kind of HDP identified were among the information collected. Additionally, each hospital’s institutional guideline for treating the spectrum of HDPs was reviewed, and with informed consent, the heads of the various labor wards filled questionnaire on the use of anti-HDP drugs. The questionnaire was about antihypertensive and anticonvulsant medication availability and their usage for treating HDPs, particularly pre-eclampsia/eclampsia. It required the names of all medicines, their dosage forms, pharmaceutical strengths, and how they were administered. Importantly, requests were made for institutional MgSO_4_ protocols and administration procedures including loading and maintenance doses, and their concentrations when given intravenously (IV) or intramuscularly (IM). Further, unit leaders answered questions on serum concentration monitoring of MgSO_4_, adverse effects and toxicity monitoring in both open and closed-ended questions.

### Ethical Approval

Permission to obtain data from the institutions in the Bono Region was granted by the Bono Regional Health Directorate, and ethical approval was sought and obtained from the Committee on Human Research Publication and Ethics of KNUST (Ref: CHRPE/AP/119/20). Authors had no access to the identities of individual participants whose data were collected, and informed consent was obtained from labor ward in-charges/doctors where questionnaires required completion.

### Data analysis

The data was analyzed using Stata 17.0 (Stata Corporation, Texas, USA). Categorical variables were compared using Chi-square (*χ*^2^) or Fisher’s exact tests, as appropriate. Binomial regression with a log-link function was used to estimate crude and adjusted odds with 95% confidence intervals (CIs) for factors associated with hypertensive disorders in pregnancy (HDPs). The dataset that supports the findings of this study has been attached under the supporting information section as S1 file_Dataset.xlsx. Missing data was excluded, and results with p-values ≤ 0.05 were considered statistically significant.

### Diagnoses of HDPs according to the standard treatment guideline of Ghana

According to Ghana’s STG, a pregnant woman has a hypertensive disorder when her blood pressure (BP) is ≥ 140/90 mmHg on two or more occasions, spaced at least five minutes apart, using an appropriate BP monitoring device (19). The diagnoses of gestational hypertension, chronic hypertension, pre-eclampsia with or without severe features, superposed pre-eclampsia and eclampsia may be made based on the timing of the onset of the hypertensive BP relative to the gestational age, time to conception, the patient’s signs and symptoms, and any biochemical assessment suggesting an organ dysfunction. Gestational hypertension is new onset, occurring after mid-gestation without proteinuria or classical signs and symptoms; chronic hypertension is similarly symptomless, carried into pregnancy or identified in the first 20 weeks of gestation, unassociated with proteinuria, and persists after labor. A non-severe and severe forms exist in pre-eclampsia. Usually, it begins during mid-gestation with a fresh onset of hypertension (SBP ≥ 140/DBP ≥ 90 mmHg) and mild proteinuria. It becomes severe if there are neurological signs and symptoms, massive proteinuria, elevated BP (SBP > 160/DBP ≥ 110 mmHg), or biomarkers related to target organ injury. Women with chronic hypertension are diagnosed with superimposed pre-eclampsia if their features align with pre-eclampsia while eclampsia is diagnosed if a pre-eclamptic woman experiences seizures without other identifiable reasons (19). It is highlighted that screening for soluble fm-like tyrosine kinase-1 (sFlt-1) and placental growth factors (PlGF) is also accessible for diagnosing HDPs in advance settings but non-existent in the region at the time of data collection.

### Pharmacotherapy of HDPs per the guidelines of Ghana

Under Ghanaian treatment policy, methyldopa and sustained release/retarded nifedipine are prescribed for mild hypertensive cases in pregnancy (BP 140/90-159/109 mmHg). The recommended treatments for severe maternal hypertension are IV hydralazine, and labetalol. To abort seizures or prevent same, MgSO_4_ by the Pritchard approach is recommended. This protocol administers the drug in two phases: an IV loading and IM maintenance doses. During loading, 14g of the anticonvulsant is administered as 4g IV in a 20% MgSO_4_ solution at first, with 2 doses of 5g IM in a 50% solution into each buttock (10g). Subsequently, series of 5g IM doses of a 50% solution are injected into alternate buttocks every four hours to initiate the maintenance phase.

This begins four hours after the loading dose has been initiated and continuously administered for 24 hours for preventive therapies or 24 hours after the last fit (19).

## Results

### Level of care of the Health Facilities involved in the study

Primary care level health services were provided by eight of the nine hospitals in rural and semi-urban communities of the region. The ninth establishment was a regional hospital that provided specialist services and doubled as the referral center for the Bono Region of Ghana.

### Parturients’ sociodemographic and obstetric data

Table 1 presents the sociodemographic data of parturients. The age range was 15-46 years and the mean age was 30.9789 (±6.80541). Those aged 26-34 years made up the largest proportion (43.9%). The majority were those who attained basic education (61.7%), working in informal occupations (68.0%), multigravida (51.9%), and multiparous women (53.0%).

**Table 1.**
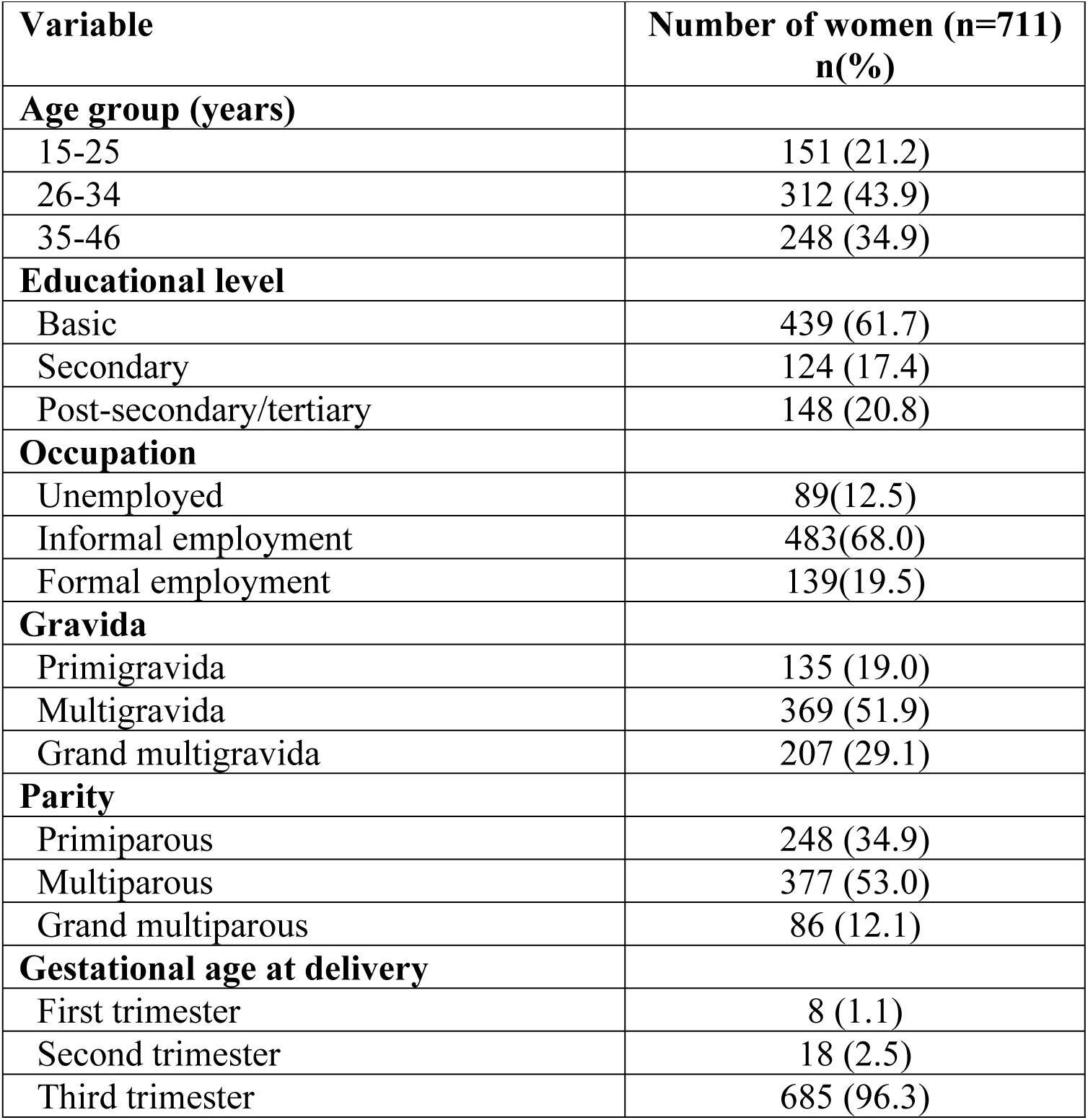
Sociodemographic data of women involved in the study.

| **Variable** | **Number of women (n=711)**<br>**n(%)** |
| --- | --- |
| **Age group (years)** |  |
| 15-25 | 151 (21.2) |
| 26-34 | 312 (43.9) |
| 35-46 | 248 (34.9) |
| **Educational level** |  |
| Basic | 439 (61.7) |
| Secondary | 124 (17.4) |
| Post-secondary/tertiary | 148 (20.8) |
| **Occupation** |  |
| Unemployed | 89(12.5) |
| Informal employment | 483(68.0) |
| Formal employment | 139(19.5) |
| **Gravida** |  |
| Primigravida | 135 (19.0) |
| Multigravida | 369 (51.9) |
| Grand multigravida | 207 (29.1) |
| **Parity** |  |
| Primiparous | 248 (34.9) |
| Multiparous | 377 (53.0) |
| Grand multiparous | 86 (12.1) |
| **Gestational age at delivery** |  |
| First trimester | 8 (1.1) |
| Second trimester | 18 (2.5) |
| Third trimester | 685 (96.3) |

### Types of hypertensive disorders diagnosed among parturients

A sum of 16,206 deliveries were conducted in the nine hospitals: 711 were diagnosed with HDPs. A prevalence of 4.4% was observed. From Fig 1, the commonest HDP among the parturients was pre-eclampsia. This occurred without severe features in 30.5% of the subjects but in 21.0%, it was associated with severe features. Gestational hypertension was found among 28.0%, whereas 6.2% developed eclampsia.

**Fig 1.**
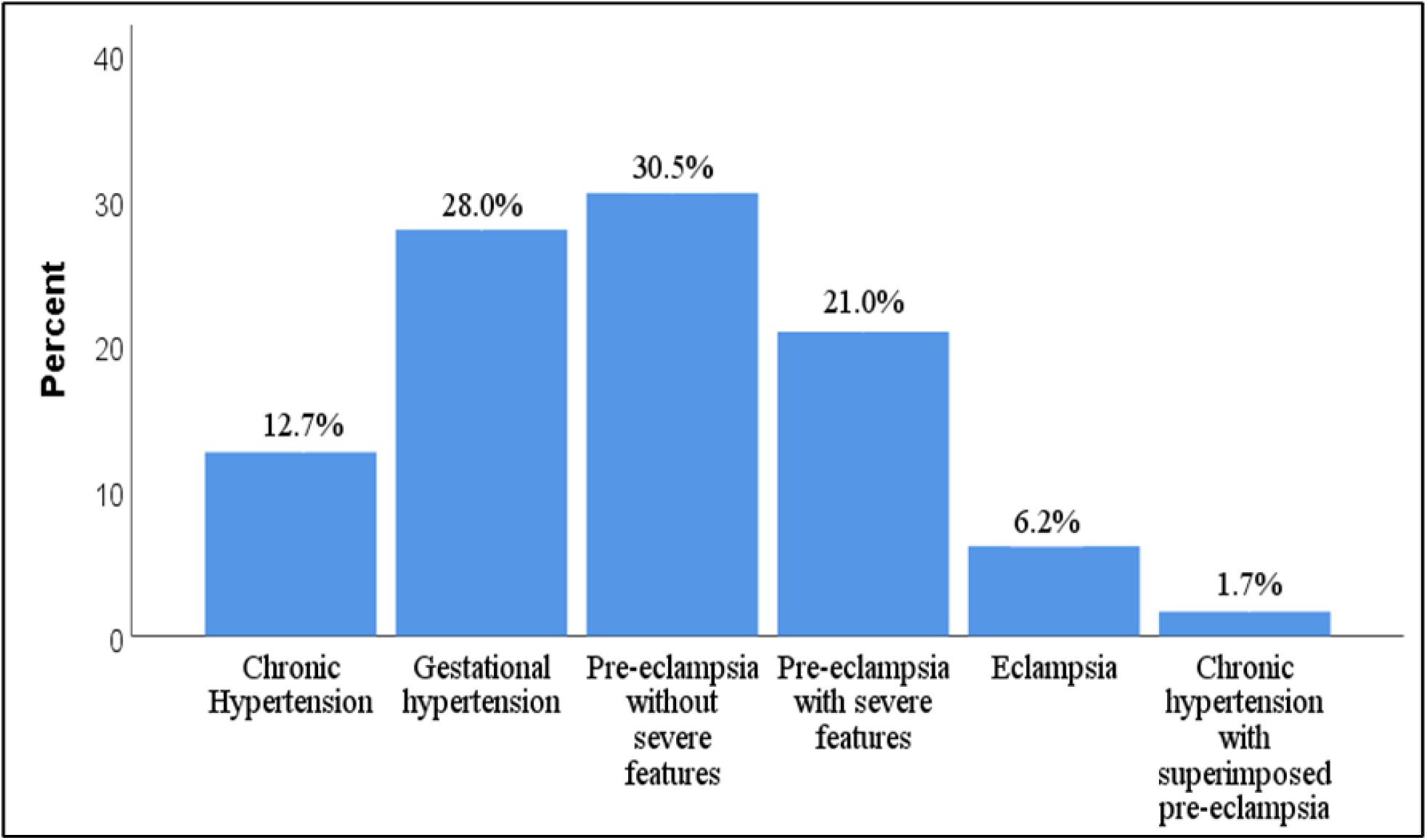
Patients’ diagnoses.

### Maternal demographic data compared with diagnoses

Table 2 summarizes the distribution of HDPs across parturients’ demographic variables. Chronic hypertension was conspicuous among women aged 35-46 years (53.3%), basic education attainers (60.0%) and multiparous women (62.2%). Gestational hypertension was prevalent among women aged 26-34 years (45.2%), the least educated (65.8%) and multiparous as well (58.8%). Pre-eclamptic disorders (non-severe, severe, and superimposed) were seen mostly in middle-aged women (46.8%) between 26-34 years, those with low literacy (59.0%), multigravida (54.2%) and multiparous (52.1%). The majority of eclamptics (68.2%) were in the 15-25 age group, the least educated (70.5%), and primiparous (77.3%) who had delivered for the first time.

**Table 2.**
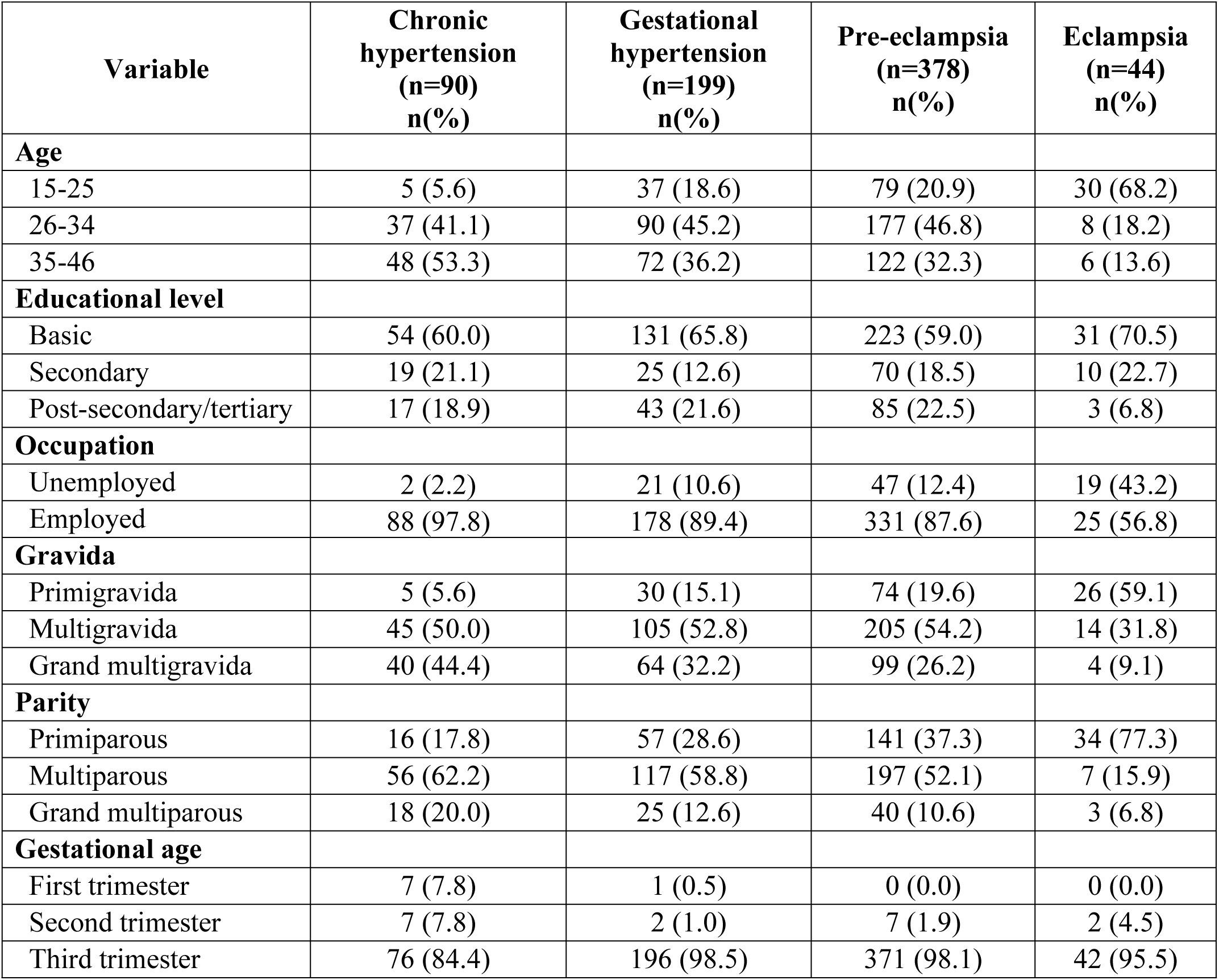
A comparison of hypertensive disorders with maternal sociodemographic characteristics.

### Factors associated with hypertensive disorders during pregnancy

The associations between parturients’ characteristics and hypertensive disorders are presented in Table 3. Age was significantly associated with hypertensive disorders during pregnancy (p<0.001). Younger women aged 15-25 years (25.8%), were observed more frequently with pre-eclampsia/eclampsia, whereas older women aged 35-46 years (41.5%), were more common in the chronic/gestational hypertension group. Maternal employment status also showed a significant association (p=0.002). A higher proportion of the unemployed (15.6%) were in the pre- eclampsia/eclampsia category, while those in employment (92.0%) dominated in the chronic/gestational hypertension group. Gravidity and parity were strongly associated with hypertensive disorders (p<0.001). Primigravida (23.7%) or primiparous women (41.5%), were more linked to pre-eclampsia/eclampsia. In contrast, grand multigravida women (36.0%) and multiparous women (59.9%) were more represented in the chronic/gestational hypertension group. However, the educational status of a parturient did not significantly impact the type of a hypertensive disorder (p = 0.411).

**Table 3.** Parturients’ factors and their association with hypertensive disorders (n=711)

| Variable | Chronic/gestational hypertension | Pre-eclampsia/eclampsia | p-value |
| --- | --- | --- | --- |
| **Age** |  |  | <0.001 |
| 15-25 | 42 (14.5) | 109 (25.8) |  |
| 26-34 | 127 (43.9) | 185 (43.8) |  |
| 35-46 | 120 (41.5) | 128 (30.3) |  |
| **Educational level** |  |  | 0.411 |
| Basic | 185 (64.0) | 254 (60.2) |  |
| Secondary | 44 (15.2) | 80 (19.0) |  |
| Post-secondary/tertiary | 60 (20.8) | 88 (20.9) |  |
| **Occupation** |  |  | 0.002 |
| Unemployed | 23 (8.0) | 66 (15.6) |  |
| Employed | 266 (92.0) | 356 (84.4) |  |
| **Gravida** |  |  | <0.001 |
| Primigravida | 35 (12.1) | 100 (23.7) |  |
| Multigravida | 150 (51.9) | 219 (51.9) |  |
| Grand multigravida | 104 (36.0) | 103 (24.4) |  |
| **Parity** |  |  | <0.001 |
| Primiparous | 73 (25.3) | 175 (41.5) |  |
| Multiparous | 173 (59.9) | 204 (48.3) |  |
| Grand multiparous | 43 (14.9) | 43 (10.2) |  |
| **Trimester of pregnancy** |  |  | 0.001* |
| First trimester | 8 (2.8) | 0 (0.0) |  |
| Second trimester | 9 (3.1) | 9 (2.1) |  |
| Third trimester | 272 (94.1) | 413 (97.9) |  |

Note: \* (Fisher's exact)

### Parturients’ odds of developing a hypertensive disorder

Table 4 examines the odds of a parturient developing pre-eclampsia/eclampsia based on sociodemographic and obstetric factors. Early maternal age (15-25 years) was 2.43 times more related to pre-eclampsia/eclampsia (cOR = 2.43; 95% CI: 1.57-3.75 p = < 0.001) compared to older parturient age (35-46 years). Unemployed parturients were twice likely to have pre- eclampsia/eclampsia (cOR = 2.14; 95% CI=1.29-3.53; p = 0.001). Primigravidity elevated the odds to almost three times that of grand multi-gravida women (cOR =2.88; 95% CI: 1.80-4.62, p < 0.001). Primiparity also doubled the odds (cOR = 2.39, 95% CI: 1.44 -3.96; p = <0.001) and after adjusting for potential confounders, primiparity remained a significant predictor though with a marginal significance level (aOR = 1.83; 95% CI: 0.97–3.46, p-value = 0.05). The educational level of a parturient did not significantly impact the odds of developing pre-eclampsia/eclampsia.

**Table 4.** Regression analysis of the association between maternal sociodemographic data with re-eclampsia/eclampsia.

| Variable | Pre-eclampsia /Eclampsia | cOR | 95% CI | p-value | aOR | 95% CI | p-value |
| --- | --- | --- | --- | --- | --- | --- | --- |
| **Age** |  |  |  | 0<0.001 |  |  | 0.37 |
| 15-25 | 109 (72.2) | 2.43 | 1.57-3.75 |  | 1.51 | 0.84-2.72 |  |
| 26-34 | 185 (59.3) | 1.36 | 0.97-1.91 |  | 1.14 | 0.78-1.67 |  |
| 35-46 | 128 (51.6) | 1 |  |  | 1 |  |  |
| **Educational level** |  |  |  | 0.40 |  |  |  |
| Basic | 254 (57.86) | 1 |  |  |  |  |  |
| Secondary | 80 (64.52) | 1.32 | 0.88-2.00 |  |  |  |  |
| Post-secondary/tertiary | 88 (59.46) | 1.07 | 0.73-1.56 |  |  |  |  |
| **Occupation** |  |  |  | 0.001 |  |  |  |
| Unemployed | 66 (74.16) | 2.14 | 1.29-3.53 |  |  |  |  |
| Employed | 356 (57.23) | 1 |  |  |  |  |  |
| **Gravida** |  |  |  | 0<0.001 |  |  |  |
| Primigravida | 100 (74.07) | 2.88 | 1.80-4.62 |  |  |  |  |
| Multigravida | 219 (59.35) | 1.47 | 1.04-2.07 |  |  |  |  |
| Grand multigravida | 103 (49.76) | 1 |  |  |  |  |  |
| **Parity** |  |  |  | 0<0.001 |  |  | 0.05 |
| Primiparous | 175 (70.56) | 2.39 | 1.44-3.96 |  | 1.83 | 0.97-3.46 |  |
| Multiparous | 204 (54.11) | 1.17 | 0.73-1.88 |  | 1.09 | 0.66-1.80 |  |
| Grand multiparous | 43 (50.00) | 1 |  |  | 1 |  |  |
| **Trimester of pregnancy** |  |  |  | 0.38 |  |  |  |
| Second trimester | 9 (50.00) | 1 |  |  |  |  |  |
| Third trimester | 413 (60.29) | 0.65 | 0.25-1.68 |  |  |  |  |

aOR, adjusted odds ratio; cOR, crude odds ratio; CI, confidence interval; p, probability

### Availability and use of antihypertensive medications for the pregnant women at the hospitals

Methyldopa and the sustained-release nifedipine were the common oral antihypertensive medicines available in all facilities for HDPs. The sustained release oral nifedipine (30mg) was used by seven hospitals as first choice agent for HDPs, particularly pre-eclampsia. Intravenous hydralazine (20mg/ml) was available and used in all the hospitals for severe cases of pre- eclampsia. Four out of nine hospitals had IV labetalol (100mg/20ml) as an alternative (Table 5).

**Table 5.** Antihypertensive medicines available in the hospitals for managing hypertension in pregnancy.

| Medicines | Number of hospitals that use the agent (n=9) | Used as first choice agent (n=9) |
| --- | --- | --- |
| **Orals** |  |  |
| Methyldopa 250mg | 9 | 2 |
| Nifedipine retard 20mg | 5 | 0 |
| Extended release nifedipine 30mg | 9 | 7 |
| Oral Hydralazine 25mg | 2 | 0 |
| Oral Labetalol 50mg/100mg | 0 | 0 |
| **Parenteral agents** |  |  |
| IV hydralazine (20mg/ml) | 9 | 9 |
| IV labetalol(100mg/20ml) | 4 | 0 |

IV, intravenous

### Usage of magnesium sulfate as anticonvulsant for pre- eclampsia/eclampsia

All the nine hospitals had stocks of MgSO_4_ injectables for managing pre-eclampsia/eclampsia. The 50% solution (10ml ampoule) was available in 8/9 facilities and protocols in the form of posters for administering same were conspicuous in all labor wards. The Pritchard regimen was the norm across facilities. Loading doses were unanimously 14g; injected as 4g IV followed by 10g IM.

Maintenance doses were 5g IM, given four hourly apart. The 50% MgSO_4_ solution was used for IM maintenance doses while IV doses were administered with a 20% solution. One hospital had in addition to the 50% protocol, another protocol that allowed a 20% solution to be used to inject 5g IM doses. Per all protocols examined, MgSO_4_ prophylactic treatment was completed 24 hours after initiating the loading and administering a series of 6 IM maintenance doses. When the loading and maintenance doses are put together, each patient receives 44g of the salt within 24 hours. Testing renal function of patients at the point of initiating MgSO_4_ therapy was not always done in any of the hospitals. All facilities lacked the capacity to monitor serum concentrations of magnesium ions in women on treatment. Therapy monitoring was solely based on patient sign/symptoms such as knee jerk responses, respiratory rate and urine output. Seven out of the nine hospitals had usable calcium gluconate injectables to manage MgSO_4_ toxicities (Table 6).

**Table 6.** Usage of MgSO_4_ to manage pre-eclampsia/eclampsia by hospitals in Bono Region.

| **MgSO<sub>4</sub> Usage** | **Facility (n=9)** |
| --- | --- |
| **Presence of MgSO<sub>4</sub> injectable at hospital** |  |
| Yes | 9 |
| **Type of stocks present at the time** |  |
| 50% MgSO<sub>4</sub> (10ml ampoule) | 8 |
| 20% MgSO<sub>4</sub> (10ml ampoule) | 1 |
| **Treatment protocol pasted at labor ward** |  |
| Yes | 9 |
| **Type of treatment regimen** |  |
| Loading dose + maintenance doses | 9 |
| **Composition of loading dose** |  |
| 4g IV + 10g IM | 9 |
| **Concentration of MgSO<sub>4</sub> by IV route** |  |
| 20% | 9 |
| 50% | 0 |
| **Maintenance doses** |  |
| 5g IM 4 hourly | 9 |
| **Concentration of MgSO<sub>4</sub> by IM route** |  |
| 50% | 8 |
| 20% | 1 |
| **Injection site for IM doses** |  |
| Buttocks | 9 |
| **Completion of treatment regimen** |  |
| After 6 IM maintenance doses injected 4 hourly | 9 |
| **Maximum dose of MgSO<sub>4</sub> administered to a patient** |  |
| 44g | 9 |
| **Renal function test results seen before starting MgSO<sub>4</sub>** |  |
| Always | 0 |
| Sometimes | 9 |
| **Patient monitoring** |  |
| Solely based on patient signs and symptoms | 9 |
| Serum concentrations of Mg<sup>2+</sup> | 0 |
| **Availability of calcium gluconate to manage toxicity** |  |
| Yes | 7 |
| No | 0 |
| Expired | 2 |

IM, Intramuscular; IV, Intravenous; MgSO<sub>4</sub>, Magnesium sulfate

## Discussion

This study highlights the prevalence of hypertensive disorders among parturients in the Bono Region of Ghana and examines how these disorders were managed at service delivery points using antihypertensive and anticonvulsant medicines. The need for the study arose from the lack of published data on HDPs in the region, which made it challenging to quantify the burden and compare same with other smaller regions of Ghana. Additionally, several years after the adoption of WHO-recommended treatment guidelines by Ghana, there have been limited post-implementation studies evaluating the administration of MgSO_4_ for pre-eclamptic/eclamptic patients. The study addresses these knowledge gaps.

Our results indicate that the prevalence of HDPs in the Bono Region was 4.4%, a rate less than those reported in Zambia (6.7%), Ethiopia (6.82%), sub-Saharan Africa (8%), and Nigeria (25.8%) (20–23). However, it was higher than Burkina Faso (1.4%) and Ghana’s Upper West Region (3.2%) (11,24). The observed regional variations could be due to differences in study methodologies, environmental and socioeconomic factors. Unlike studies that collected data from antenatal clinics (ANCs) attendees, our study focused on data from women in labor, potentially influencing prevalence estimates. Hospital deliveries in most Ghanaian facilities usually rank lower than ANC attendance which may contribute to differences in findings (25,26). This may be due to factors such as transportation challenges, financial constraints, fear of cesarean section, and some negative perceptions of hospital staff attitudes which could contribute to home deliveries in rural settings, leading to an underestimation of HDP prevalence in hospitals (25,26). However, in Burkina Faso and the Upper West Region of Ghana, where the occurrences were lower than our data, it seems that the effects of attendance and social characteristics were more prominent (11,24).

Pre-eclampsia was the most prevalent HDP in this study, consistent with findings of Dassah et al. (2019) in Kumasi, Ghana (13). Among affected women, 30.5% had non-severe pre-eclampsia while one-fifth had pre-eclampsia with severe features. These rates exceed the prevalence (4.1%) estimated in a systematic review of sub-Saharan Africa indicating that our findings may reflect regional differences in healthcare access and resources (22). Previous researchers showed that HDPs were predicted by maternal age (27,28). Our multivariable regression analysis revealed a significant association between age and HDP. Younger parturients (15-25 years) were more prone, particularly to pre-eclampsia/eclampsia. Decreasing maternal age increased the odds, with over two-thirds of eclamptic cases occurring in this age group (see Table 2). This trend is supported by previous studies in Ghana and South Africa (29,30). The pathophysiological basis for this association remains unclear but may involve mothers’ aberrant immune responses to paternally inherited fetal antigens during first pregnancies as hypothesized in earlier literature which could apply to younger women getting pregnant for the first time (31). Additionally, reduced ANC attendance among teenage mothers due to social stigma may lead to poor pregnancy monitoring, anemia, and inadequate preventive care, further increasing their risk (28). Contrarily, our advanced-aged parturients (≥ 35 years) were more aligned with chronic hypertension which was consistent with existing literature (20,32,33). This is conceivable, given that obesity, vascular calcification, stiffness, and loss of distensibility correlate with old age and greatly impact hypertension (34).

The results of this study relates to previous authors who reported that parity plays a role in pre- eclampsia/eclampsia (28,35). While our crude odds ratio indicated a significant association between primigravidity and HDPs (p < 0.001), the adjusted odds showed significance for primiparity (p = 0.05). Known in literature, multigravidity has been associated with a more regulated immune response and adaptation to repeated exposure to paternally inherited antigens as opposed to primigravid/primiparous women, who are first time encounters, less adapted and therefore prone to inflammatory response leading to pre-eclampsia (31). Whereas parity was a significant predictor, educational level did not show an association. Unlike some prior studies, our findings did not establish a significant relationship between HDPs and illiteracy after adjusting for other variables (27,28). The lack of variability in educational attainment among study participants may have limited the statistical power to detect such an association.

Maternal hypertensives were primarily treated with the 30mg extended release nifedipine, 250mg methyldopa, and IV hydralazine (20 mg/ml). These agents were consistently available across all the nine hospitals. Among the available oral agents, nifedipine was the most commonly prescribed first-line treatment in seven out of nine hospitals, mirroring prescribing patterns observed at Tamale Teaching Hospital in Ghana (36). Its preference over methyldopa may be attributed to its faster onset of action (30-45 minutes vs. 1-1.5 hours for methyldopa) (37–39). Oral labetalol was rarely stocked, so its use for treatment was limited, likely due to cost and availability. Nifedipine’s use was in line with Ghana’s treatment policy, but some evidence suggests that nifedipine may increase the risk of disease progression to pre-eclampsia when used in non-severe maternal hypertension, prompting WHO to advocate for alternative oral agent such as labetalol in non- severe hypertensive cases (40).

Some recently revised guidelines and review publications appear to place IV hydralazine as second line agent for severe pre-eclampsia due to its perceived unfavorable kinetics and side effects, but its utilization as first choice agent was evident in this study (41). The widespread usage was also observation in Nigerian (42). The high utilization of IV hydralazine, despite recent guidelines favoring IV labetalol may be driven by staff familiarity, affordability, and availability (43).

All study sites adhered to national and international guidelines recommending MgSO₄ for managing severe pre-eclampsia/eclampsia (19,44). All hospitals had access to the product, along with its administration protocols. The Pritchard regimen was uniformly followed, likely due to its convenience and independence from infusion pumps, which are often scarce in resource-limited settings (19). Magnesium sulfate is administered to severe pre-eclamptic women not because they have Mg^2+^ deficiency, but rather to raise Mg^2+^ plasma concentrations to counteract Ca^2+-^-induced muscle twitches, vasospasms, and glutamate-mediated neuronal excitement in eclamptic and severely pre-eclamptic women (45–47). Moreover, the goal is to hinder cholinergic transmission at the neuromuscular junction to stop or avoid convulsive episodes (46,47).

Under Pritchard method, patients receive an initial loading dose of 14g MgSO₄. This is implemented as 4g slow IV infusion using a 20% solution, which provides immediate anticonvulsant activity lasting about 30 minutes. This is followed by a 10g intramuscular (IM) injection using a 50% solution, extending the anticonvulsant effect for 3-4 hours. To maintain therapeutic levels, a 5g IM dose is injected into alternating gluteal muscles every 4 hours for up to 24 hours (46,47). Eight hospitals stocked the 50% w/v MgSO₄ in 10 ml ampoules in compliance with WHO requirements (48). However, one hospital only had 20% w/v MgSO₄, presenting a significant clinical challenge. A 5g IM dose of 20% MgSO₄ requires 25 ml, exceeding the regular injection volume of 10 ml for gluteal muscles from a 50% solution (19,49). Some hospital pharmacists might have underestimated the health implication of substituting a 50% w/v MgSO₄ solution with a 20% for IM doses. Such an excess volume may cause pain, drug leakage, inflammation, and abscess formation (50,51). This issue, as highlighted by Babu et al. (2022) may contribute to poor patient compliance with IM maintenance therapy (52).

At the sites, severe pre-eclamptic/eclamptics received six IM maintenance doses based on the MgSO_4_ protocols evaluated. These amounted to cumulative doses of 44g MgSO_4_ for each patient in 24 hours as opposed to 40g reported in some literature (53). Despite high adherence to the MgSO₄ guidelines, none of the hospitals had the capacity to monitor serum Mg²⁺ levels, relying instead on urine output, respiratory rate, and patellar reflexes, a practice that may be insufficient for preventing toxicity. Given the risk of accumulation in patients with impaired renal function, dose adjustments based on weight or alternative regimens like the Dhaka regimen or a 12-hour protocol proposed by Beyuo et al. ( 2022) may be safer options for facilities without laboratory capacity (54–56).

The study had some drawbacks. For example, part of the data was collected by observing drug protocols, drug shelves, and medicine trays. In addition, procedural data were gathered from physicians, labor/maternity unit heads, and pharmacists rather than by watching actual procedures being performed which might differ. Moreover, inaccurate records might have been captured in labor registers, which could impact the quality of the data collected. Again, the study did not include health centers, private clinics, or maternity homes since it was assumed that many HDP patients would be directed to major hospitals because such smaller facilities were unable to handle maternal complications or conduct caesarean surgeries when urgent deliveries were required. If some HDPs were managed without referral, this would have an effect on the statistics that this study projects.

## Conclusions

The prevalence of HDP in the Bono Region of Ghana was 4.4%. Pre-eclampsia was the most common HDP. Young parturient age, unemployment, primigravida and primiparity were the predictors of pre-eclampsia/eclampsia. The extended-release oral nifedipine and IV hydralazine were the main therapies for HDPs, especially for severe pre-eclamptic patients. Magnesium sulfate protocols for pre-eclampsia/eclampsia conformed to standard regimen. The supply of 20% MgSO_4_ solutions for IM maintenance doses was inappropriate due to its larger injection volumes that may be harmful to patients. Standardizing the concentration of magnesium sulfate solutions for pre-eclampsia/eclampsia could optimize intramuscular dosing under Pritchard regimen, improving treatment consistency.

## Acknowledgments

We appreciate the Bono Regional Health Directorate and the medical directors or administrators of all the hospitals for granting us permission to access hospital records for data. We also appreciate the cooperation of midwives, doctors and pharmacists in the hospitals for providing some information that was needed for this study.

## Supporting information

**S1 Fig. Patients’ diagnoses**

**S1 Table. Sociodemographic data of women involved in the study**

**S2 Table. A comparison of hypertensive disorders with maternal sociodemographic characteristics**

**S3 Table. Parturients’ factors and their association with hypertensive disorders (n=711)**.

*Note: * (Fisher’s exact)*

**S4 Table. Regression analysis of the association between maternal sociodemographic data with pre-eclampsia/eclampsia**. aOR, adjusted odds ratio; cOR, crude odds ratio; CI, confidence interval; p, probability

**S5 Table. Antihypertensive medicines available in the hospitals for managing hypertension in pregnancy**. IV, intravenous

**S6 Table. Usage of MgSO_4_ to manage pre-eclampsia/eclampsia by hospitals in Bono Region**. IM, Intramuscular; IV, Intravenous; MgSO_4_, Magnesium sulfate

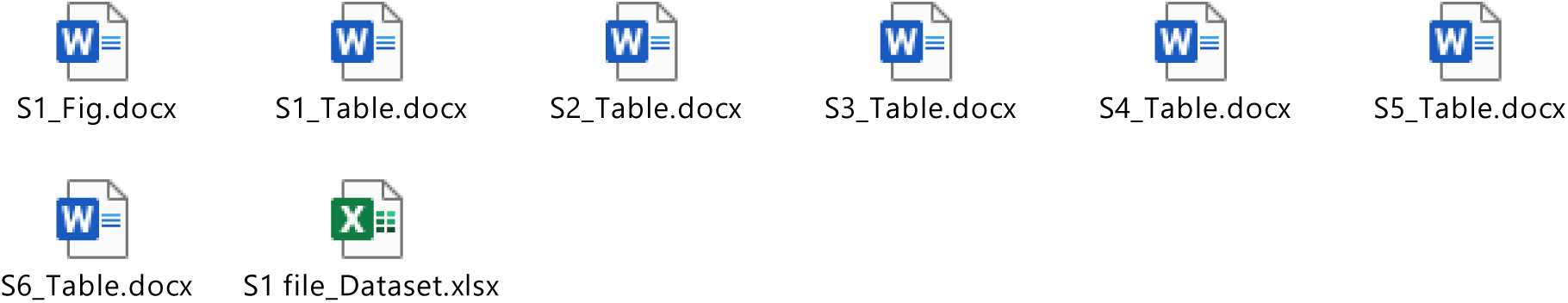

## Data availability statement

All relevant data are within the manuscript and its supporting information file attached

## Authors’ contributions

**Conceptualization**: Francis Fordjour, Edward Tieru Dassah and Kwame Ohene Buabeng

**Data curation**: Francis Fordjour, Bernard Okyere, Kwadwo Addai-Darko

**Data analysis**: Francis Fordjour, Jonathan Boakye-Yiadom and Edward Tieru Dassah

**Methodology**: Francis Fordjour, Edward Tieru Dassah and Kwame Ohene Buabeng,

**Mentorship & supervision**: Edward Tieru Dassah and Kwame Ohene Buabeng,

**Writing-original draft**: Francis Fordjour, Bernard Okyere, Jonathan Boakye-Yiadom and Edward Tieru Dassah

**Writing-review & editing:** Kwadwo Addai-Darko, Edward Tieru Dassah, and Kwame Ohene Buabeng

